# Utilization of Eight Antenatal Care Contacts Among Adolescent and Adult Mothers at a Referral Hospital in Western Kenya: A Comparative Cross-Sectional Study

**DOI:** 10.1101/2025.09.14.25335701

**Authors:** Reuben Nyongesa Kere, Jackton Omoto, Patrick Nyamohanga Marwa

## Abstract

**Introduction:** The World Health Organization (WHO) recommends eight antenatal care (ANC) contacts. Kenya adopted this new model in 2022, yet adherence remains low. This study investigated factors influencing the frequency of eight ANC contacts among adolescent and adult mothers in Kenya, using Andersen and Newman’s Behavioural Model for Health Services Utilization.

**Methods:** We conducted a comparative analytical cross-sectional study at Jaramogi Oginga Odinga Teaching and Referral Hospital (JOOTRH), Kisumu, Kenya, involving a stratified sample of 73 adolescents and 219 adult mothers. We collected data using a questionnaire and the Mother and Child Health Handbook. We used descriptive statistics, the Mann-Whitney U test, chi-square test and multivariable logistic regression analysis.

**Results:** Adolescent mothers were less likely to have insurance coverage (74%) compared to adult mothers (90.9%) (p = 0.001) and reported distance to the ANC facility as a barrier (42.5%, p = 0.016). Adherence to eight contacts was low (10.5% for adult mothers and 8.2% for adolescent mothers), with median ANC contacts of 5.00 IQR 3-6 and 4.00 IQR 3-5.5, respectively. Mothers with high-risk pregnancies (AOR = 2.3281, 95% CI: 1.4691-3.9108, p = 0.005) were more likely to complete eight ANC contacts.

**Conclusion:** The low adherence rates highlight a critical gap in care for non-high-risk pregnancies, whose low compliance appears driven by a reduced perceived need. Health systems must revise antenatal clinical pathways to increase the perceived value of frequent contacts for all mothers and integrate financial support, such as health insurance enrolment, to address economic barriers, especially for adolescents.

## Introduction

Maternal morbidity and mortality are global public health concerns, with preventable factors being the cause of over 70% of maternal fatalities. Sub-Saharan Africa (SSA) bears a share of this burden, accounting for approximately 90% of deaths in 2023 [1].

Antenatal care (ANC) is universally recognized as the foundation of safe motherhood. It offers an environment for health promotion, the prompt detection and management of complications and readiness for childbirth and related emergencies [2, 3]. Consistently utilizing ANC services decreases adverse pregnancy outcomes [4]. To achieve better maternal and neonatal outcomes, the World Health Organization (WHO) updated its 2016 guidelines, transitioning from a four-visit model to eight or more ANC contacts [3, 5].

Despite these established advantages, adherence to the eight-contact model remains low across many low- and middle-income countries (LMICs) [6]. In Kenya, for instance, only a small fraction of mothers (4%) complete the recommended eight ANC contacts, indicating a significant break in the continuity of maternal care [7].

This study employed Andersen and Newman’s Behavioural Model for Health Services Utilization as its theoretical basis, adapting it to the new eight-contact ANC model (as shown in Figure 1) [8]. This enables analysis of how predisposing, enabling, and need factors influence the frequency of the eight ANC contacts [9]. Prior research confirms that maternal education, socioeconomic status, and place of residence influence ANC attendance [10, 11]. However, a key void in knowledge remains concerning the specific predisposing, enabling, and need variables that drive the completion of eight ANC contacts under the new model in the Kenyan setting [12, 13, 14]. In addition, most existing local research focused on the four-visit model in different regional settings, which limits its relevance to the current eight-contact model in Western Kenya [15].

**Figure 1.**
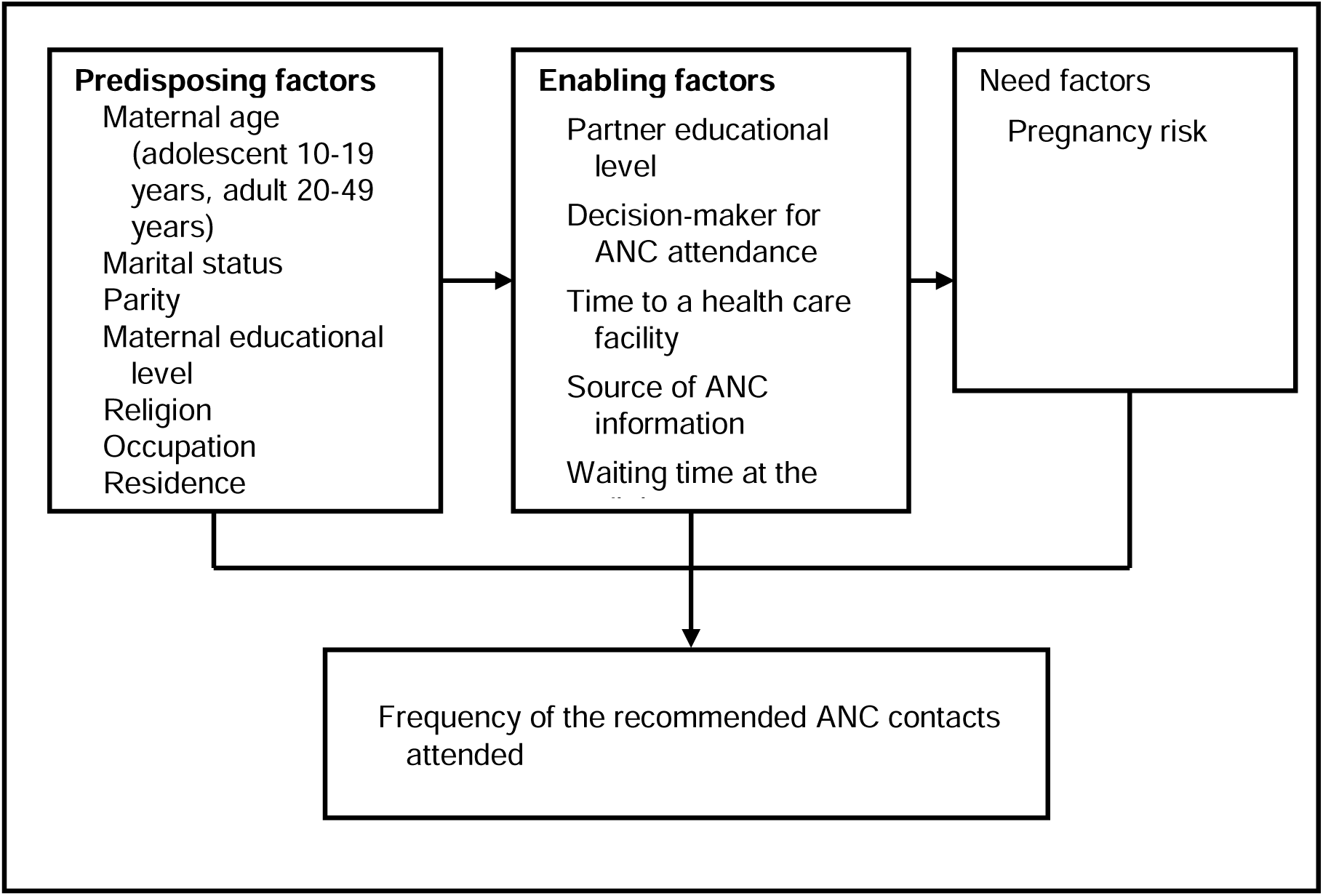
Theoretical Framework (Andersen & Newman, 2005; Andersen, 2008)

To fill this key gap, we conducted a comparative analysis of the factors affecting the frequency of the recommended eight ANC contacts among adolescent and adult mothers at JOOTRH, thereby providing valuable, age-specific data to inform targeted policy interventions.

## Methodology

### Study design, site and setting

We conducted a comparative, analytical cross-sectional study at Jaramogi Oginga Odinga Teaching and Referral Hospital (JOOTRH) in Kisumu, Kenya, from April to July 2025. The study investigated differences in the frequency of eight Antenatal Care (ANC) contacts between adolescent mothers (15–19 years) and adult mothers (20–49 years). Data collection took place in the maternity department to maximize maternal recall and allow for verification of ANC records using the Mother and Child Health Handbook.

### Study population

All mothers admitted to the maternity department at JOOTRH.

### Selection criteria

Postpartum mothers (adolescent: 15–19 years; adult: 20–49 years) admitted to the JOOTRH maternity department were included with informed consent. We excluded mothers who were clinically unstable or lacked a Mother and Child Health Handbook, which was essential for data validation.

### Sample size calculation

We calculated the sample size to achieve sufficient statistical power to detect significant differences in the frequency of eight antenatal contacts between adolescent and adult mothers. The outcome was the number of the recommended eight ANC contacts attended. We used a two-population proportion formula: n= (Zα/2+Zβ) 2∗ (p1 (1−p1) +p2 (1−p2))/ (p1−p2)2. The assumed proportion of mothers was 19.4% (P1) for adolescents and 37.5% (P2) for adults. We derived these numbers from previous studies [10, 16], respectively. To account for a smaller expected difference in the new eight-contact model and a 10% non-response/incomplete record, we used a total sample size of n=292. Based on the maternity department’s admission ratio, we applied a 3:1 ratio (adult to adolescent). This targeted 73 adolescent mothers and 219 adult mothers to maximize statistical efficiency for the comparative analyses.

### Sampling method

We used a stratified random sampling technique, grouping participants into two groups: adolescent mothers (15-19 years) and adult mothers (20-49 years). This stratification ensured adequate representation of both groups and reduced sampling error. Within each stratum, we used random sampling to select participants. To meet the required sample size (n=292), participants were recruited using a 3:1 ratio based on the maternity department’s admission ratio of adult to adolescent mothers.

### Conceptual framework

This study uses and adapts Andersen and Newman’s Behavioural Model for Health Services Utilization (Figure 1). This demonstrates that predisposing, enabling, and need factors influence adherence to the eight recommended ANC contacts.

## Operational definitions of variables

### Dependent variable

Frequency of the recommended ANC contacts: The total number of ANC contacts a pregnant woman attended, as documented in her Kenya Mother and Child Health Handbook, compared to the WHO 2016 recommended eight or more contacts.

### Independent variables

Predisposing factors: Maternal age: A participant’s age in years, categorized as adolescent (15-19 years) or adult (20-49 years). Marital status: A participant’s current legal marital status. Parity: The number of times a participant has delivered after 28 weeks’ gestational age. Maternal educational level: The level of formal education a participant has completed. Religion: A participant has self-reported religious affiliation. Enabling factors: Occupation: A participant’s primary type of work. Residence: A participant’s usual place of residence. Partner educational level: The highest level of education completed by the participant’s partner. Decision-maker for ANC attendance: The person or group who primarily influences the participant’s decision to attend ANC services. Distance to a healthcare facility: A participant’s ease of reaching the nearest healthcare facility offering ANC services. Waiting time at the clinic: The time in hours that a participant spends at the clinic for their ANC appointment. Source of ANC information: The primary source from which the pregnant woman received information about ANC. Healthcare insurance cover: Whether a participant had health insurance that covers ANC services. Need factors: Pregnancy risk: The likelihood of obstetrical complications affecting maternal or fetal health during pregnancy. We scored this using the Modified Coopland Antenatal Risk Scoring System [17].

### Quantitative variables

Maternal age groupings: Adolescent (15–19 years) and Adult (20–49 years). We categorized the maternal age variable for a comparative analysis. ANC contacts groupings: The final multivariable analysis used a binary grouping to compare optimal adherence (8 or more contacts) against all forms of suboptimal adherence (1–7 contacts). Suboptimal adherence represented all contacts falling short of the current WHO 2016 recommendation for a minimum of eight ANC contacts. For other quantitative variables, we categorized them into distinct groupings.

### Data Collection Tools and Procedures

We collected data using a questionnaire and a Mother and Child Health Handbook review. The handbook provided the objective ANC data on the frequency of antenatal contacts. The questionnaire, developed in English and Kiswahili, captured predisposing, enabling, and need factors, including pregnancy risk categorized by the Modified Coopland antenatal risk scoring system [17]. We refined the questionnaire following a pilot test on a 5% sample of each age group to ensure clarity and effectiveness.

### Measures to minimize bias

We managed selection bias through stratified random sampling to ensure population representativeness. We minimized recall bias by using a structured, pre-tested questionnaire and the Mother and Child Health Handbook corroboration. Finally, we controlled for confounding bias in the analysis by applying a multivariable logistic regression model.

### Data management and analysis

We collected data using the Open Data Kit (ODK) app, uploaded it into the OnaData database and coded it to ensure participant anonymity during analysis. We restricted access to the authorized research team. We analyzed data using IBM SPSS Statistics (version 26.0). We used descriptive statistics, the Mann-Whitney U test and the Chi-square test to compare the frequency of ANC contacts between the two age groups. Multivariable logistic regression analysis identified associated factors (AORs and 95% CIs). P-value of <0.05 was significant.

### Ethical considerations

The study adhered to the ethical principles outlined in the Declaration of Helsinki. We obtained informed consent from all participants after explaining the study objectives. In Kenya, an adolescent mother who has delivered is an emancipated minor, legally allowing her to provide valid consent for research [18]. We maintained confidentiality and anonymity, stored data securely and restricted access. Maseno University gave initial approval, followed by ethical approval (ISERC/JOOTRH/129/24) from the JOOTRH Institutional Scientific and Ethical Review Committee. The National Commission for Science, Technology and Innovation (NACOSTI) granted a research permit (NACOSTI/P/25 /417736) on April 09, 2025.

## Results

The final study sample included 292 participants, 73 (25%) adolescent mothers (aged 15-19 years) and 219 (75%) adult mothers (aged 20-49 years).

### Predisposing factors

Analysis of predisposing factors (Table 1) found a strong association between maternal age and marital status (p<0.001): the majority of adolescent mothers (69.9%) were single, while most adult mothers were married (77.2%). Parity also differed significantly (p<0.001). Almost all adolescent mothers (97.3%) had one or two pregnancies, while a substantial proportion of adults (43.4%) had three or more. Education level was significantly associated with maternal age (p<0.001) (Table 1). Adolescent mothers were more likely to have secondary education (54.8%), while adult mothers were more likely to have post-secondary education (50.2%).

**Table 1:** Predisposing Factors of adolescent and adult mothers at JOOTRH.

|  |  |  |  |  |  |  |  |
| --- | --- | --- | --- | --- | --- | --- | --- |
| Independent variable | Category | Adolescent mothers<br>N (%) | Adult mothers<br>N (%) | Total<br>N=292 | Chi-Square<br>($\chi^2$) | df | P value |
| Marital Status | Single | 51(69.9) | 46(21.0) | 97(33.2) | 61.231 | 3 | <0.001 |
|  | Married | 20(27.4) | 169(77.2) | 189(64.7) |  |  |  |
|  | Divorced/separated | 2(2.7) | 3(1.4) | 5(1.7) |  |  |  |
|  | Widowed | 0(0.0) | 1(0.5) | 1(0.3) |  |  |  |
| Parity | 1-2 | 71(97.3) | 124(56.6) | 195(66.8) | 40.828 | 2 | <0.001 |
|  | 3-4 | 2(2.7) | 72(32.9) | 74(25.3) |  |  |  |
|  | 5+ | 0(0.0) | 23(10.5) | 23(7.9) |  |  |  |
| Level of education | Primary school | 13(17.8) | 55(25.1) | 68(23.3) | 23.112 | 2 | <0.001 |
|  | Secondary school | 40(54.8) | 54(24.7) | 94(32.2) |  |  |  |
|  | Post-secondary school | 20(27.4) | 110(50.2) | 130(44.5) |  |  |  |
| Religion | Christian | 72(98.6) | 214(97.7) | 286(97.9) | 0.227 | 1 | 0.634 |
|  | Muslim | 1(1.4) | 5(2.3) | 6(2.1) |  |  |  |

### Enabling factors

Analysis of enabling factors (Table 2) showed a strong association between maternal age and occupation (p<0.001). A majority of adolescent mothers were students (68.5%), and most adult mothers were unemployed (76.3%). This difference extended to their partners’ occupation (p<0.001); adolescent mothers’ partners were more often students or unemployed. This instability contributed to almost all adolescent mothers (97.3%) reporting poor household income (p<0.001). Prominent geographical barriers were also found, with a significant difference in residence (p<0.001). Most adolescent mothers (78.1%) resided in rural areas, and 42.5% of them reported that distance was a problem (p=0.016) compared to adult mothers (27.4%). In addition, health insurance coverage differed significantly (p=0.001), with adolescents being much more likely to lack any coverage (26%) compared to adult mothers (9.1%). Critically, the source of ANC information showed a significant difference (p<0.001). Adolescents predominantly relied on informal networks like family and friends (87.7%) (Table 2), while adult mothers reported getting more information from healthcare providers (44.7%).

**Table 2:** Enabling Factors of adolescent and adult mothers at JOOTRH.

| Table 2: Enabling Factors of adolescent and adult mothers at JOOTRH |  |  |  |  |  |  |  |
| --- | --- | --- | --- | --- | --- | --- | --- |
| Independent Variable | Category | Adolescent Mothers N (%) | Adult Mothers N (%) | Total N=292 | Chi-Square ($\chi^2$) | df | P value |
| Current occupation | Employed | 0 (0.0) | 17 (7.8) | 17 (5.8) | 74.379 | 2 | <0.001 |
|  | Unemployed | 23 (31.5) | 167 (76.3) | 190 (65.1) |  |  |  |
|  | Student | 50 (68.5) | 35 (16.0) | 85 (29.1) |  |  |  |
| Occupation of partner | Employed | 4 (5.5) | 53 (24.2) | 57 (19.5) | 48.299 | 2 | <0.001 |
|  | Unemployed | 38 (52.1) | 147 (67.1) | 185 (63.4) |  |  |  |
|  | Student | 31 (42.5) | 19 (8.7) | 50 (17.1) |  |  |  |
| Monthly household income | Poor | 71 (97.3) | 168 (76.7) | 239 (81.8) | 15.560 | 1 | <0.001 |
|  | Middle | 2 (2.7) | 51 (23.3) | 53 (18.2) |  |  |  |
| Residence | Urban | 16 (21.9) | 109 (49.8) | 125 (42.8) | 17.350 | 1 | <0.001 |
|  | Rural | 57 (78.1) | 110 (50.2) | 167 (57.2) |  |  |  |
| ANC Attendance decision marker | Self | 22 (30.1) | 94 (42.9) | 116 (39.7) | 3.739 | 2 | 0.154 |
|  | Partner | 4 (5.5) | 10 (4.6) | 14 (4.8) |  |  |  |
|  | Others | 47 (64.4) | 115 (52.5) | 162 (55.5) |  |  |  |
| Distance to ANC facility | No problem | 42 (57.5) | 159 (72.6) | 201 (68.8) | 5.795 | 1 | 0.016 |
|  | Big problem | 31 (42.5) | 60 (27.4) | 91 (31.2) |  |  |  |
| Waiting time at the ANC clinic | 0–1 hr | 34 (46.6) | 94 (42.9) | 128 (43.8) | 1.654 | 2 | 0.437 |
|  | 2–3 hrs | 38 (52.1) | 115 (52.5) | 153 (52.4) |  |  |  |
|  | 4+ hrs. | 1 (1.4) | 10 (4.6) | 11 (3.8) |  |  |  |
| Partner's Educational Level | No formal | 1 (1.4) | 1 (0.5) | 2 (0.7) | 3.294 | 3 | 0.348 |
|  | Primary | 10 (13.7) | 39 (17.8) | 49 (16.8) |  |  |  |
|  | Secondary | 15 (20.5) | 29 (13.2) | 44 (15.1) |  |  |  |
|  | Postsecondary | 47 (64.4) | 150 (68.5) | 197 (67.5) |  |  |  |
| Health insurance type | Government | 53 (72.6) | 191 (87.2) | 244 (83.6) | 14.026 | 2 | 0.001 |
|  | Private | 1 (1.4) | 8 (3.7) | 9 (3.1) |  |  |  |
|  | None | 19 (26.0) | 20 (9.1) | 39 (13.4) |  |  |  |
| ANC information source | Healthcare provider | 8 (11.0) | 98 (44.7) | 106 (36.3) | 32.571 | 2 | <0.001 |
|  | Media | 1 (1.4) | 12 (5.5) | 13 (4.5) |  |  |  |
|  | Others | 64 (87.7) | 109 (49.8) | 173 (59.2) |  |  |  |

### Need factors

They played a differentiating role when examining previous history (Table 3). Adult mothers had a statistically higher occurrence of several previous obstetrical and gynaecological conditions: previous low birth weight or macrosomia (p=0.001), previous caesarean section (p=0.003), stillbirth or neonatal death (p=0.021), and a difficult or prolonged labour (p=0.041). These findings suggest that adult mothers were more likely to have a complex previous obstetrical history. However, the Modified Coopland Antenatal Risk Score distribution showed no statistically significant difference between the two groups overall (p=0.612). Finally, the comparison of early ANC initiation (≤12 weeks) between adults (37%) and adolescents (26%) was not significant (p=0.087).

**Table 3:** Previous Obstetrical and Gynaecological Conditions.

| Table 3: Previous Obstetrical and Gynaecological Conditions |  |  |  |  |  |  |  |
| --- | --- | --- | --- | --- | --- | --- | --- |
| Independent Variable | Category | Adolescent Mothers N (%) | Adult mothers N (%) | Total N=292 | Chi-Square ($\chi^2$) | df | P value |
| Infertility | No<br>Yes | 15(100.0)<br>0(0.0) | 157(99.4)<br>1(0.9) | 172(99.4)<br>1(0.6) | 0.671 | 1 | 0.413 |
| 2 or more first-trimester abortions | No<br>Yes | 14(93.3)<br>1(6.7) | 152(96.2)<br>6(3.8) | 166(96.0)<br>7(4.0) | 0.955 | 1 | 0.328 |
| Second-trimester abortions | No<br>Yes | 13(86.7)<br>2(13.3) | 150(94.9)<br>8(5.1) | 163(94.2)<br>10(5.8) | 0.671 | 1 | 0.413 |
| Child birth <2.5kg or >4kg | No<br>Yes | 13(86.7)<br>2(13.3) | 129(81.6)<br>29(18.4) | 142(82.1)<br>31(17.9) | 10.716 | 1 | 0.001 |
| Previous caesarean section | No<br>Yes | 11(73.3)<br>4(26.7) | 126(79.7)<br>32(20.3) | 137(79.2)<br>36(20.8) | 8.902 | 1 | 0.003 |
| PPH or manual removal of placenta | No<br>Yes | 14(93.3)<br>1(6.7) | 146(92.4)<br>12(7.6) | 160(92.5)<br>13(7.5) | 3.519 | 1 | 0.061 |
| Previous stillbirth or neonatal death | No<br>Yes | 14(93.3)<br>1(6.7) | 143(90.5)<br>15(9.5) | 157(90.8)<br>16(9.2) | 5.309 | 1 | 0.021 |
| Prolonged/difficult labour | No<br>Yes | 15(100.0)<br>0(0.0) | 149(94.3)<br>9(5.7) | 164(94.8)<br>9(5.2) | 4.171 | 1 | 0.041 |
| Gestational hypertension/preeclampsia | No<br>Yes | 13(86.7)<br>2(13.3) | 143(90.5)<br>15(9.5) | 156(90.2)<br>17(9.8) | 3.540 | 1 | 0.060 |
| Eclampsia | No<br>Yes | 15(100.0)<br>0(0.0) | 158(100.0)<br>0(0.0) | 173(100.0) | - | - | - |
| Gestational diabetes | No<br>Yes | 14(93.3)<br>1(6.7) | 157(99.4)<br>1(0.6) | 171(98.1)<br>2(1.2) | 0.671 | 1 | 0.413 |

### Frequency of ANC Contacts

The Mann-Whitney U test indicated a significant difference in the total number of ANC contacts between groups (p=0.049). Adult mothers achieved a higher median of 5.00 contacts (Interquartile Range [IQR]: 3.0–6.00) compared to adolescent mothers’ 4.00 contacts (IQR: 3.0–5.5) (Table 4).

**Table 4:** Measures of central tendency and Mann-Whitney U test of the total number of recommended antenatal contacts between adolescent (10-19 years) and adult mothers (20-49 years) at JOOTRH.

| Independent variable | Category | Number (%) | Median | IQR | U-statistic | P value |
| --- | --- | --- | --- | --- | --- | --- |
| Frequency of ANC contacts | Adolescent mothers (15-49 years) | 73 (25) | 4.00 | 3.0 – 5.5 | 8020.0 | 0.0498 |
|  | Adult mothers (20-49 years) | 219 (75) | 5.00 | 3.0 – 6.0 |  |  |

Despite this difference in the distribution of the total count, a Chi-square test on the categorized ANC frequency (1–4, 5–7, 8+ contacts) yielded no significant difference (p=0.468) (Table 5). This difference likely arises from the χ2 test losing statistical power by collapsing the continuous ANC count into broad categorical groups. The largest proportion of adolescent mothers (53.4%) completed only 1–4 contacts, while the largest proportion of adult mothers (44.3%) completed 5–7 contacts. Furthermore, adults had a higher rate of completing the recommended eight or more contacts, 10.5% compared to 8.2%.

**Table 5:** Chi-square test of the frequency of recommended antenatal contacts between adolescent (15-19 years) and adult mothers (20-49 years).

|  |  |  |  |  |  |  |  |
| --- | --- | --- | --- | --- | --- | --- | --- |
| Independent variable | Category | Adolescent mothers N (%) | Adult mothers N (%) | Total N=292 | Chi-Square ($\chi^2$) | df | P value |
| Frequency of recommended antenatal care contacts | 1-4 | 39(53.4) | 99(45.2) | 138(47.3) | 1.521 | 2 | 0.468 |
|  | 5-7 | 28(38.4) | 97(44.3) | 125(42.8) |  |  |  |
|  | 8+ | 6(8.2) | 23(10.5) | 29(9.9) |  |  |  |

### Multivariable analysis of factors associated with ANC contact frequency

After adjusting for predisposing and enabling factors, the only significant predictor in the multivariable logistic regression (Table 6) was pregnancy risk category. Mothers with high-risk pregnancies (AOR=2.3281, 95% CI: 1.4691−3.9108, p=0.005) were more likely to attend the recommended number of contacts. We must interpret this finding within the context of the referral setting.

**Table 6:** Logistic regression table of factors associated with the frequency of the recommended antenatal care contacts among adolescent (15-19 years) and adult mothers (20-49 years).

|  |  |  |  |  |
| --- | --- | --- | --- | --- |
| Independent variable | Category | AOR | 95% CI for AOR | P value |
| Age | Adult mothers (Ref.) |  |  |  |
|  | Adolescent mothers | 0.8519 | (0.7137-1.3952) | 0.2757 |
| Marital Status | Single (Ref.) |  |  |  |
|  | Married | 2.0732 | (0.8959-4.9187) | 0.0819 |
|  | Divorced | 1.9622 | (0.0519-8.8078) | 0.6971 |
| Parity | 1-2 (Ref.) |  |  |  |
|  | 3-4 | 0.7934 | (0.3387-1.8623) | 0.5929 |
|  | 5+ | 0.5587 | (0.1448-2.0948) | 0.3892 |
| Education | Primary (Ref.) |  |  |  |
|  | Secondary | 0.8048 | (0.3453-1.8716) | 0.6131 |
|  | Post-secondary | 0.6704 | (0.2680-1.6484) | 0.3861 |
| Occupation | Employed (Ref.) |  |  |  |
|  | Unemployed | 0.2437 | (0.0113-1.8425) | 0.2363 |
|  | Student | 0.2123 | (0.0087-2.1285) | 0.2345 |
| Residence | Urban (Ref.) |  |  |  |
|  | Rural | 1.4807 | (0.7267-3.0607) | 0.2829 |
| ANC Attendance decision maker | Self (Ref.) |  |  |  |
|  | Partner | 1.6073 | (0.3946-7.2951) | 0.5149 |
|  | Others | 1.1866 | (0.6294-2.2431) | 0.5966 |
| Risk Category | Low risk (Ref.) |  |  |  |
|  | Moderate risk | 1.2652 | (0.5116-1.4563) | 0.2507 |
|  | High risk | 2.3281 | (1.4691-3.9108) | 0.0054 |
| Health insurance type | Government type (Ref.) |  |  |  |
|  | Private | 1.0696 | (0.1397-8.0672) | 0.9468 |
|  | None | 0.6364 | (0.2672-1.4775) | 0.2974 |
| Partner Occupation | Employed (Ref.) |  |  |  |
|  | Unemployed | 1.0767 | (0.3098-3.8604) | 0.9071 |
|  | Student | 0.5145 | (0.1169-2.2679) | 0.3747 |
| Information about ANC | Others (Ref.) |  |  |  |
|  | Media | 0.7505 | (0.2401-1.5144) | 0.3843 |
|  | Healthcare providers | 0.7064 | (0.3389-1.4608) | 0.3494 |
| Income | Poor (Ref.) |  |  |  |
|  | Middle | 1.1725 | (0.3094-4.7162) | 0.8172 |
| Distance | No problem (Ref.) |  |  |  |
|  | Big problem | 0.9548 | (0.4830-1.8941) | 0.8943 |

Note: Ref. =Reference category, AOR=Adjusted Odds Ratio, CI=Confidence Interval

## Discussion

### Statement of Principal Findings

Our study applied Andersen and Newman’s Behavioural Model for Health Services Utilization [8, 9] to examine factors influencing the use of eight antenatal care contacts. The most compelling finding was the significant association between high-risk pregnancy status and adherence. Mothers identified as high-risk were 2.3 times more likely to attend all eight recommended ANC contacts (AOR = 2.3281; p=0.005). This result confirms the success of the ’need’ component of the Andersen-Newman model [8, 9]. It validates the critical role of the Modified Coopland Antenatal Risk Scoring System and demonstrates the effectiveness of the clinical referral and monitoring system in enforcing adherence for those with the highest medical need [17].

However, the low adherence among low-risk and moderate-risk groups suggests a systemic failure related to the perceived value of frequent contacts. We observed initial differences between the groups: Adult mothers had a higher median of 5.00 ANC contacts compared to 4.00 for adolescents. Adolescent mothers were also significantly more likely to lack any health insurance coverage (26%) compared to adult mothers (9.1%). However, our multivariable analysis showed that these access barriers did not predict adherence in the final model. The urgency of a high-risk diagnosis appears to override both the enabling factors (distance, cost) and the predisposing factors (age, marital status, education). For adolescent and adult mothers who were high-risk, strong motivation compelled them to complete the eight contacts.

This suggests that the health system at JOOTRH is highly effective at mobilizing antenatal care when clinical need is clear. We must consider the study’s location, a national referral hospital. Mothers reaching JOOTRH form a pre-selected group because either they overcame initial access problems or the system referred them due to complex antenatal needs. The clinical need for high-risk care simply became the dominant factor, masking the underlying issues that affect low-risk adolescent and adult mothers differently. This contrast reveals a fundamental problem. Clinical need successfully overcomes the challenges faced by mothers of all ages.

Therefore, while high-risk adherence is a positive indicator, the challenge remains in redesigning services to enhance the perceived value and accessibility for all non-high-risk mothers. The healthcare system must create an equally strong perceived benefit for the majority of mothers who are not high-risk. We must design services that motivate low-risk adolescent and adult mothers to value and complete the full eight contacts.

### Strengths and Weaknesses of the Study

The primary strength of this study is its contribution to addressing a critical knowledge gap in the Kenyan context by investigating factors influencing the frequency of the new WHO eight-contact ANC model. The comparative cross-sectional design was a key methodological strength; it allowed for a direct comparison of ANC utilization patterns between adolescent and adult mothers at a major national referral hospital, enhancing the study’s ability to identify specific differences. The finding of a relationship between pregnancy risk and ANC contact frequency represents a significant contribution. This key finding shows a gap in care for mothers with low and moderate-risk pregnancies. They did not see the need to achieve the recommended number of ANC contacts because of their perceived low need for antenatal care, unlike mothers with high-risk pregnancies.

This study has two limitations that future research could address. Firstly, this was a cross-sectional study; this prevents us from establishing a causal relationship between variables. Additionally, this study reported adherence rates to the eight contacts of 8.2% for adolescents and 10.5% for adult mothers. These rates are higher than the documented national adherence rate of 4% in Kenya [8]. This difference is attributable to the study’s specific setting, which attracts mothers with high-risk pregnancies who have successfully navigated the lower levels of antenatal care or a more health-aware patient population. This introduces a potential selection bias, limiting the generalizability of these rates to primary healthcare facilities across Western Kenya. Future research should target primary care settings to obtain a more representative measure of national adherence under the new WHO model.

### Strengths and Weaknesses in Relation to Other Studies

The suboptimal utilization of the eight-contact schedule observed in our study (8.2% and 10.5% completion for adolescents and adults, respectively) is broadly consistent with the low national adherence rate of 4% in Kenya [7]. However, our completion rates are considerably lower than those documented in other regional studies, specifically the 18.35% reported in Ethiopia [20] and 23.4% in Uganda [21]. A comprehensive meta-analysis across low- and middle-income settings established the pooled utilization of eight or more ANC visits at 18.11%, placing the challenge identified locally towards the lower end of the global spectrum for service uptake [6].

The tendency for adolescent mothers to discontinue ANC attendance aligns with existing findings from Uganda and a systematic review, which cited factors such as stigma, inadequate social support, and fragmented service coordination as primary obstacles [22, 23]. Additionally, our results concur with a multilevel analysis of African countries, which proposes that socioeconomic impediments and irregular support structures compromise follow-through [24]. In line with research from Ethiopia, our findings support the idea that prior interactions with the healthcare system impact subsequent attendance decisions [25].

The observed demographic variance between our age groups likely shapes their perception and understanding of ANC, a pattern echoed in studies conducted throughout East Africa [26]. This distinction directly relates to the financial impediment we identified, the significant difference in health insurance coverage, an issue also noted by studies in Ethiopia [27].

### Discussion of Important Differences in Results

Our study revealed clear divergences in predisposing, enabling, and need components between the two maternal age groups. Most adolescent mothers were either nulliparous or of low parity, generally possessing a secondary education. In contrast, adult mothers exhibited varied parity, with a substantial number having more than two pregnancies, alongside a higher incidence of post-secondary education attainment. These educational and parity differences are likely to shape their health literacy, awareness, and perceptions regarding ANC utilization, a pattern observed across East Africa [26].

Socioeconomic status also demonstrated a marked contrast in occupational profile. The majority of adolescents were students, whereas most adult mothers reported being unemployed. For adolescents, being students, often coupled with partners who were also unemployed or students, signifies considerable financial vulnerability, contributing to the nearly universal poverty reported by this demographic. This financial precarity directly translates into an access barrier, specifically evidenced by the significantly lower health insurance coverage among adolescents, an issue also noted by Ethiopian studies [27].

The vital differentiating factor centred on the mothers’ need’ variables, particularly their past obstetric records, which acted as a powerful determinant for engaging in subsequent ANC attendance. Adult mothers reported a statistically greater incidence of a complicated obstetric history, encompassing conditions like prior caesarean sections, low birth weight, stillbirth or neonatal death, or difficult/prolonged labour, a conclusion supported by a recent cohort study in Rwanda [28]. This cumulative experience is hypothesized to intensify their perceived necessity for consistent ANC attendance in current pregnancies, often overriding common logistical hurdles. This finding confirms that a complex obstetrical history strongly influences care-seeking in multiparous women, consistent with findings in Rwanda on the effect of previous adverse outcomes on care-seeking behaviour [28].

In contrast, the adolescent mothers in our study generally lacked a complicated obstetrical past. The absence of previous adverse outcomes often resulted in their classification as low-risk pregnancies. Although clinical standards frequently categorize adolescent pregnancies as high-risk due to combined biological and social factors [5], the reliance of a risk stratification tool on past obstetric events leads to a potential underestimation of risk in our adolescent sample [29]. A study in Kenya has also highlighted this limitation, noting that current risk stratification tools fail to account for the unique social and biological risks inherent to adolescents [30]. This resulting low-risk designation inadvertently leads to a reduced perceived need for frequent ANC contacts among adolescents, as their uncomplicated pregnancy course may lead them to question the value of additional contacts. A previous Kenyan study links the severity of pregnancy complications to higher ANC attendance rates, supporting this idea [19]. Therefore, a major difference exists: while past complications motivate care seeking in adult mothers, the absence of such a history may unknowingly lead to underutilization of frequent ANC contacts among adolescents, consistent with observations from Ethiopia [31].

### Meaning of the Study

The observed correlation between a heightened perception of antenatal risk and greater ANC contact frequency provides strong affirmation for the ’need factors’ element of the Newman and Anderson model [8, 9]. This essentially means that women conscious of greater risk in their pregnancy are more compelled to comply with medical guidance, a conclusion supported by parallel research in Malaysia and a large multi-country investigation spanning Africa and Asia [32, 33].

Notably, the absence of a link between ANC adherence and predisposing and enabling determinants indicates that the perceived health risk of the current pregnancy serves as a more powerful stimulus for consistent attendance than a mother’s socioeconomic or demographic profile within our specialized referral setting.

Nonetheless, the observed rate of antenatal contact discontinuation among adolescents draws attention to specific hurdles, including a lack of autonomy and health insurance coverage. This necessitates the creation of specialized targeted service delivery methods. Given the striking finding regarding the lack of health insurance among adolescents, incorporating health financing initiatives directly into maternal care services is deemed crucial for fostering greater adherence in this vulnerable group.

### Unanswered Questions and Future Research

While we found that mothers with low and moderate-risk pregnancies underutilize the recommended number of ANC contacts due to a lower perceived need, the underlying reasons for this perception remain an unanswered question [31]. Future qualitative research should explore the specific beliefs of mothers with low risk and moderate risk regarding the importance of the eight ANC contacts. The finding that a high-risk diagnosis in pregnancy can both motivate attendance and that the same complications that define risk act as barriers requires further studies.

Longitudinal studies should establish the causal relationship between a high-risk diagnosis, health system support, and the final number of ANC contacts achieved. A study in Ghana showed that with a dedicated follow-up, adolescent mothers achieved a similar or even higher frequency of ANC contacts than their adult counterparts [34]. Future research should evaluate such dedicated follow-up interventions within the Kenyan health system to determine their feasibility and effectiveness in closing the utilization gap of eight ANC contacts for adolescent mothers.

## Conclusion

Adherence to the new eight-contact WHO Antenatal Care (ANC) model remains low for both adolescent and adult mothers, though this is significantly higher among those with high-risk pregnancies. This pattern suggests the current system is highly responsive to the ’need’ component of the Andersen-Newman model, where perceived necessity is the primary driver, overshadowing predisposing or enabling factors. This difference highlights a crucial need to re-evaluate ANC service delivery and address barriers for low- and moderate-risk mothers, particularly the low perceived value of frequent contacts and inadequate health insurance coverage among adolescents. To improve adherence, health systems must integrate supportive financial mechanisms, such as mandatory insurance enrolment. In addition, it is essential to revise antenatal clinical pathways. Low- and moderate-risk mothers require revised pathways—perhaps featuring fewer, more comprehensive group sessions—to increase the perceived value and engagement necessary for completing all eight contacts. Finally, future research should target primary healthcare facilities to gain a more accurate understanding of how predisposing and enabling factors truly affect adherence.

### What is already known on this topic

- Globally, adherence to the new WHO eight-contact ANC model is a significant challenge, particularly in LMIC.
- Disparities in ANC attendance exist between adolescent and adult mothers, with adolescents often having fewer contacts due to socio-economic challenges.
- Higher maternal education and financial stability are generally associated with better ANC attendance.

### What this study adds

- This study provides a comparative analysis of the utilization of the eight ANC contacts under the new eight-contact model, filling a gap in the literature focused on the Kenyan context.
- It highlights a key finding that mothers with high-risk pregnancies are significantly more likely to attend the recommended number of ANC contacts compared to those with low and moderate-risk pregnancies, due to their perceived need for antenatal care.
- Adolescent mothers demonstrate significantly greater financial barriers, evidenced by a higher likelihood of lacking health insurance coverage compared to adult mothers, contributing to their high rate of drop-off after initial ANC contacts.

## Data Availability

All data produced in the present work are contained in the manuscript

## Acknowledgements

The authors wish to acknowledge the Department of Obstetrics and Gynaecology at Maseno University, especially Dr. Gwer S.O., Dr. Muruka K., and Dr. Wameyo A., for their guidance and support. We thank the administration, staff of JOOTRH, and the postnatal ward staff for their collaboration and support. We are deeply grateful to the participants. The authors declare that no funding supported the research, authorship, and/or publication of this article.

## Authors’ Contributions

Reuben Nyongesa Kere: Led the conception and design, managed data collection, analysis and interpretation, and wrote the manuscript. Jackton Omoto: Contributed to the conception and design, and reviewed the manuscript. Patrick Nyamohanga Marwa: Contributed to the conception and design, and reviewed the manuscript. All authors read and approved the final manuscript.

